# Certified nursing assistants’ perspectives on their role in advance care planning for older persons: a qualitative study

**DOI:** 10.64898/2026.01.27.26344835

**Authors:** Patricia Jepma, Annigje AE Bos, Oumaima Boulahfa, Corine HM Latour, Bianca M Buurman, Marjon van Rijn

**Author notes:** **Corresponding author:** Patricia Jepma, Department of Medicine for Older People, Amsterdam UMC, de Boelelaan 1109, 1081, HV Amsterdam, the Netherlands.

## Abstract

**Background:** Advance care planning in older persons with palliative care needs is often not (timely) initiated. Certified nursing assistants are closely involved in the (daily) care for older persons and have important insights regarding their clients’ care preferences and quality of life. However, their role in advance care planning is currently overlooked.

**Aim:** To examine the perspectives of nursing assistants on their role in advance care planning for older persons.

**Design:** A qualitative descriptive study design using semi-structured interviews.

**Setting/participants:** Fifteen nursing assistants working in community care and nursing homes in the Netherlands were interviewed between March and December 2023. A combined inductive and deductive thematic analysis was performed using the Capability, Opportunity, Motivation Behavioral model.

**Results:** Most nursing assistants were not structurally involved in advance care planning and were in need of additional knowledge and skills (capability). The team culture affected nursing assistants’ opportunities and motivation to participate in advance care planning both positively and negatively. The opportunity was further shaped by nursing assistants’ relationship with clients and relatives and time management, while motivation also depended on personal circumstances.

**Conclusion:** This study found several barriers and facilitators affecting nursing assistants’ role in advance care planning for older persons. Structurally involving nursing assistants in advance care planning, supporting their work environment, and empowering them can foster equal collaboration with other healthcare professionals. This might contribute to the timely initiation of advance care planning and palliative care for older persons.

**Key statements:** *What is already known about the topic?:*

- Advance care planning enables patients, relatives, and involved healthcare professionals to define and discuss goals and preferences for current and future medical treatment and care.
- Certified nursing assistants are often closely involved in the (daily) care for older persons and their relatives, providing important insights into their preferences and quality of life.
- Nursing assistants are currently not structurally involved in the interprofessional collaboration regarding advance care planning.

*What this paper adds:*

- Variation was observed in nursing assistants’ roles in advance care planning across care teams and settings, with most indicating a need for additional knowledge and skills (capability).
- The team culture affected nursing assistants’ opportunities and motivation to participate in advance care planning both positively and negatively.
- The opportunity was further shaped by nursing assistants’ relationship with clients and relatives and time management, while motivation was also depended on personal circumstances.

*Implications for practice:*

- Structural involvement of certified nursing assistants in advance care planning can contribute to timely initiation of advance care planning and palliative care for older persons.
- A supportive work environment that enables nursing assistants to develop their skills and grow professionally can facilitate an advance care planning process involving the expertise of all healthcare professionals.
- The empowerment of nursing assistants could help them to become a more equal partner in advance care planning in collaboration with other involved professionals.

## Introduction

The number of older persons with care needs will increase in the coming decades. Factors such as multimorbidity, including life-limiting and chronic progressive diseases, recurrent hospital admissions, frailty and functional decline may indicate a need for palliative care.^1-4^ Advance care planning, as an integral part of palliative care, can contribute to timely conversations between older persons, relatives and healthcare professionals to define and discuss goals and preferences for current and future care and medical treatment.^5^ However, advance care planning in older persons is currently often initiated too late or not initiated at all, as potential palliative care needs are often not timely recognized.^6, 7^

In the Netherlands, nursing assistants are one of the largest professional groups in the care for older persons (with potential palliative care needs), both in nursing homes and in community care. They follow a 3-year practice-oriented vocational education program to be able to autonomously provide practical daily care, including support in activities of daily living, observation of (changing) care needs, administering medication, and performing interventions such as wound care.^8, 9^ Worldwide, there is considerable variation in the terminology used to refer to these professionals (such as certified nursing assistant, personal care assistant, or healthcare assistant), as well as in their educational preparation.^10^ They all have in common that these professionals observe care preferences and key aspects regarding the quality of life.^11^ While final responsibility for palliative care rests with the general practitioner or a medical specialist, nursing assistants’ insights can provide valuable insights for identifying palliative care needs and supporting advance care planning conversations. However, nursing professionals are currently not structurally and formally involved.^12, 13^ Common challenges and barriers have internationally been reported such as a lack of knowledge, skills and self-efficacy, unclear roles and responsibilities regarding interprofessional collaboration, and difficulties in communication with clients and relatives.^12-16^ Most of these studies focus on registered nurses, resulting in fewer insights into the role and perspectives of nursing assistants on their involvement in advance care planning.

The awareness of the importance of interprofessional advance care planning for older persons, including the role of nursing professionals, is rising.^13, 17, 18^ However, the role of nursing assistants and their perspectives on their involvement in advance care planning in the Netherlands are currently overlooked. In-depth insights into their unique perspectives on barriers and facilitators within the Dutch context may provide valuable lessons for better understanding and supporting similar healthcare professionals internationally. This may help clarify the role of nursing assistants in developing and implementing future interprofessional advance care planning interventions. Therefore, this study examines nursing assistants’ perspectives on their role in advance care planning.

## Methods

### Research question

What role do nursing assistants have or want to have in advance care planning for older persons, and what barriers and facilitators do they experience?

### Design

This study used a qualitative descriptive approach to gain an in-depth understanding of nursing assistants’ roles and perspectives regarding advance care planning for older persons.^19^ This study is reported following the Consolidated Reporting of Qualitative Research checklist.^20^

### Setting

This study explored nursing’ assistants role in advance care planning for older persons in different care settings. Nursing assistants in nursing homes worked on both somatic and psychogeriatric wards, whereas those in community care often supported a mixed population with both types of conditions. Their organisations were affiliated with the University Network of Organisations for Care for Older Adults (UNO Amsterdam), a network of 24 care organiations in the western and central Netherlands linked to the Amsterdam University of Medical Centers.

The Dutch palliative care context, including the current role of nursing assistants, is further described in supplementary file 1.

### Population

Nursing assistants were eligible if they worked in a nursing home or in community care and if they had experience with caring for patients in the palliative phase. No exclusion criteria were applied.

### Sample

Purposive sampling was used to strive for variation in age, years of work experience and setting. Additional participants were subsequently recruited via snowballing. The sample size was guided using the concept of information power, considering the study aim, sample specificity, theoretical framework, response quality, and analytic strategy.^21^ Data collection continued until no new insights relevant to the research aim emerged.

### Recruitment

Participants were recruited via contact persons of organisations affiliated with the XXX network, flyers in nursing team offices, and an online newsletter. Interested participants received study information by email, and provided written informed consent prior to an in-person interview.

### Data collection

Interviews took place between March 2023 and December 2023 at a location chosen by the participant or online via Microsoft Teams. The first two interviews were pilot interviews and conducted by two of the three researchers (OB and PJ or AB and PJ), who were all female. The following interviews were conducted by only one researcher (OB or AB). All were trained in qualitative research. AB (MA) and PJ (PhD) are registered nurses and have experience with qualitative research regarding advance care planning by nurses.

The Capability, Opportunity, Motivation Behavioral model was used to develop the semi-structured interview guide, as it helped explore nursing assistants’ roles and experiences in advance care planning.^22^ *Capability* refers to their psychological and physical capacity to engage in advance care planning. *Opportunity* captures external and environmental factors influencing their role. *Motivation* encompasses the (un)conscious processes that shape their habits, emotional reactions, and decision-making regarding taking a role in advance care planning. The capability and opportunity components also affect the motivation. Furthermore, these three components interact to influence actual behavior, providing a framework to understand the roles nursing assistants have and want to have in advance care planning for older persons, as well as the barriers and facilitators they experience.

Data collection and analysis of the interviews were performed iteratively, which meant that the researchers moved back and forth between sampling, data collection, and analysis. All interviews were audio recorded and lasted 45-60 minutes. Neither researcher had any previous relationships with the included participants.

### Data analysis

All interviews were transcribed ad verbatim. Then, thematic analysis was performed using an inductive and deductive approach (MAXQDA 2022). Inductive thematic analysis allows themes to be identified from the data, while the Capability, Opportunity, Motivation Behavioral model was used for deductive analysis based on preconceived themes.^23^ Initially, open coding of the first five interviews was performed by OB and discussed with PJ. Thereafter, axial coding was used for the remaining interviews by OB and AB separately to reorganize and cluster the codes into the behavioral model and additional concepts that were identified from the data. Lastly, the codes were further defined and structured, leading to a final coding framework. Then, corresponding quotes were selected, the research question was answered, and the findings were compared with the literature.

### Ethical issues

The Medical Ethics Review Committee at Amsterdam University Medical Center approved our interviews (approval: 2022.0868) on January 25, 2023.

## Results

Fifteen nursing assistants from seven different organisations were interviewed. All were female and aged between 23 and 61 years (mean 52 years, SD 10). Nine participants worked in community care, and six participants worked in nursing homes. Their years of experience varied between 0.5 year to 39 years (median 20 years, IQR 5-34 years) (table 1). The Capability, Opportunity, Motivation Behavioral model resulted in seven subthemes that represented nursing assistants’ capability, opportunity and motivation in advance care planning for older persons (table 2).

**Table 1.**
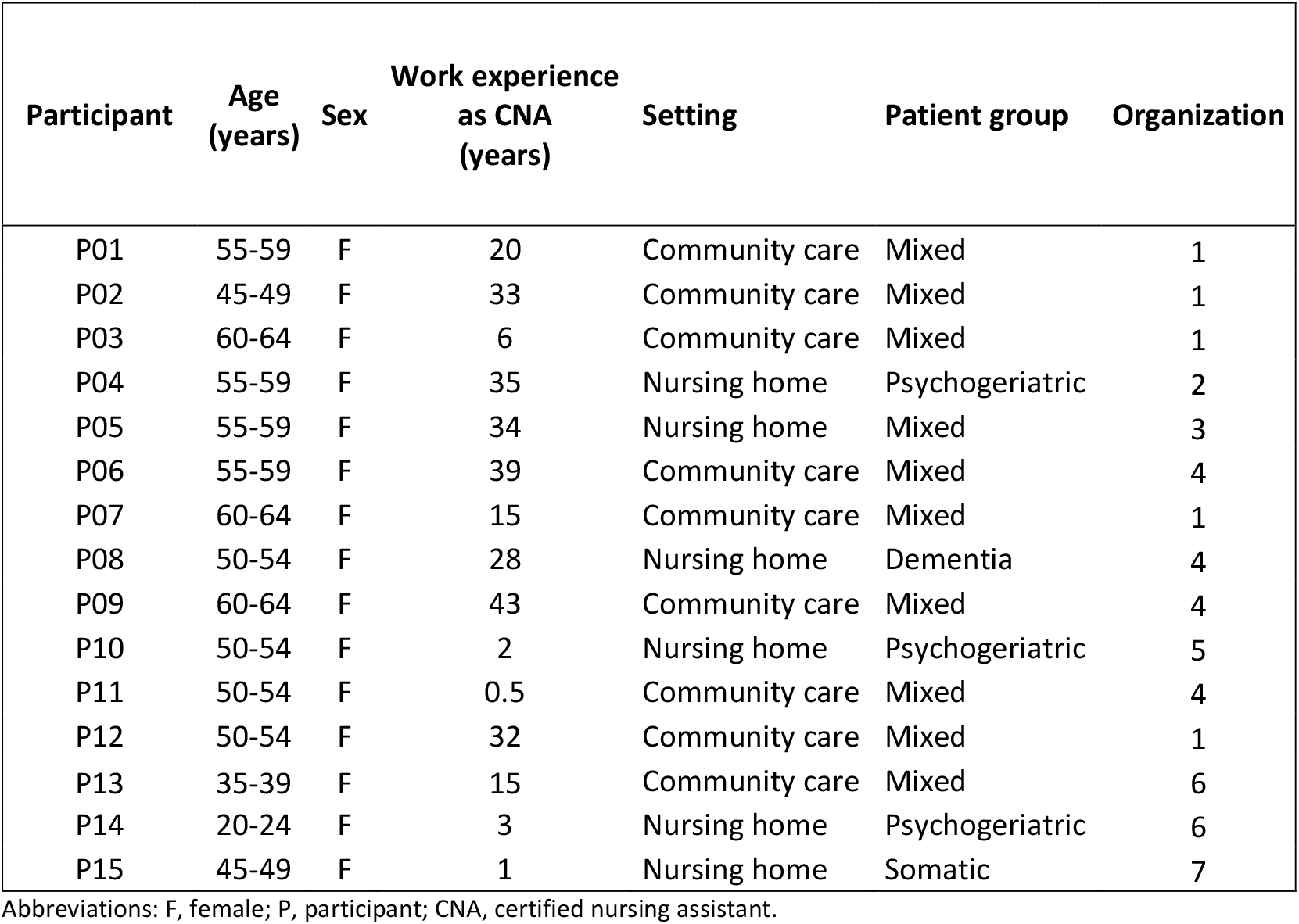
Participant characteristics.

| Participant | Age (years) | Sex | Work experience as CNA (years) | Setting | Patient group | Organization |
| --- | --- | --- | --- | --- | --- | --- |
| P01 | 55-59 | F | 20 | Community care | Mixed | 1 |
| P02 | 45-49 | F | 33 | Community care | Mixed | 1 |
| P03 | 60-64 | F | 6 | Community care | Mixed | 1 |
| P04 | 55-59 | F | 35 | Nursing home | Psychogeriatric | 2 |
| P05 | 55-59 | F | 34 | Nursing home | Mixed | 3 |
| P06 | 55-59 | F | 39 | Community care | Mixed | 4 |
| P07 | 60-64 | F | 15 | Community care | Mixed | 1 |
| P08 | 50-54 | F | 28 | Nursing home | Dementia | 4 |
| P09 | 60-64 | F | 43 | Community care | Mixed | 4 |
| P10 | 50-54 | F | 2 | Nursing home | Psychogeriatric | 5 |
| P11 | 50-54 | F | 0.5 | Community care | Mixed | 4 |
| P12 | 50-54 | F | 32 | Community care | Mixed | 1 |
| P13 | 35-39 | F | 15 | Community care | Mixed | 6 |
| P14 | 20-24 | F | 3 | Nursing home | Psychogeriatric | 6 |
| P15 | 45-49 | F | 1 | Nursing home | Somatic | 7 |

Abbreviations: F, female; P, participant; CNA, certified nursing assistant.

**Table 2.** Identified themes and subthemes.

| Main themes | Subthemes |
| --- | --- |
| Capability | Varying roles in advance care planning<br>Perceived lack of knowledge and skills |
| Opportunity | Relationship of trust with clients and relatives<br>Opportunities within the care team<br>Time pressure |
| Motivation | Personal factors as (de)motivator<br>The influence of team culture |

### Capability

#### Varying roles in advance care planning

At the start of the interview, the majority of the nursing assistants were not familiar with the term advance care planning. After the explanation, many recognized the concept and mentioned terms as ‘thinking ahead’, ‘care planning’ and ‘delivering care according to clients’ wishes’.

Variation was observed in nursing assistants’ current role in advance care planning between care teams and care settings. In nursing homes, advance care planning was mostly a task of the elderly care physician or advanced nurse practitioner. Some nursing assistants were the first point of contact for a client and revised goals and wishes minimal two times a year, although this was mostly related to daily care tasks instead of advance care planning and palliative care. Nursing assistants in community care worked more independently. Their role in advance care planning varied, depending on the opportunities within their team and organisation. Most nursing assistants minimally mentioned an observing role in clients’ health and (palliative) care needs, and these observations were subsequently shared with other involved professionals:

> *“I think we are always proactive. Only there was never a name tag on it. (…) That’s how we already work. And especially in community care you are proactive, because you have to think about a lot of things at the same time. (…) You are the eyes and ears for the nurses and the general practitioner. So you’re actually a bit of a spider in everyone’s web*.*” (P11)*

#### Perceived lack of knowledge and skills

The majority of the participants considered it important to have communication skills, for example, asking the right questions, not using difficult medical terms, being empathetic, and listening. Most nursing assistants were not educated in advance care planning during their basic training. They therefore felt insecure, for example, about their communication skills in burdensome conversations, and in interprofessional collaboration:

> *“Because I find that quite loaded and confrontational conversations. When I think about my own training as a nursing assistant, very little attention was paid to palliative care. So I definitely think it would be good if that is a larger part of the education*.*”* (P07)
>
> *“I’m unsure about that [collaboration with a general practitioner] (*..*). I don’t really know how things work with general practitioners. There was a case that really went way over my head. (…) So then the community nurse helped me. (…) Yes, then I am just a nursing assistant, and I feel that*.*”* (P13)

Participants mentioned the need for additional training in palliative care and communication skills. Current facilitators for enhancing their skills and professional growth in advance care planning included discussions during team meetings and feedback from colleagues. Furthermore, some participants mentioned that their prior work and life experiences positively contributed to their ability to engage in advance care planning:

> *“I find such conversations no problem at all. I think this is also due to my age and my years of experience in healthcare. I can imagine that if you are younger and just starting out in healthcare, these kinds of conversations are a bit more difficult*.*” (P08)*

### Opportunity

#### Relationship of trust with clients and relatives

Most of the nursing assistants emphasized the importance of building a relationship of trust with the client and relatives. They first created a safe environment by listening, taking their time and making clients feel that they were being taken seriously. Most nursing assistants mentioned that informal advance care planning conversations regarding clients’ care and treatment preferences were often not planned, but arose spontaneously during care moments like bathing and dressing:

> *“Sometimes if you take a quiet moment and take care of activities of daily living, it is a very intimate moment. You are washing people and that also results in intimate conversations*.*” (P11)*

Nursing assistants experienced that clients and relatives preferred them to be present during formal advance care planning conversations (e.g., with the general practitioner or registered nurse). Nursing assistants interpreted this as a result of the trust they had earned from clients and their families, as well as their deep understanding of the clients’ needs.

> *“The family also doesn’t know a coordinating nurse like that at all. (…) They always see us when they visit their father or mother. They can always come to us with their questions and at a certain point you build up a relationship with certain families. So yes, families do like it better when we are there*.*” (P08)*

However, nursing assistants mentioned that opportunities to discuss advance care planning depended on clients’ readiness. This influenced the way nursing assistants were able to provide care according to clients’ preferences:

> *“Some people don’t want to talk about it. They say, ‘It’s my life’. That is none of your business*.*” You will then have to respect that [*…*] The disadvantage is that you get less access or contact with someone. If that resident suddenly becomes ill and does not want to share anything, then that conversation is almost impossible*.*” (P05)*

#### Opportunities within the care team

Nursing assistants reported that team leaders, team collaboration and team culture were important factors influencing their opportunity for professional development, including advance care planning. Participants described team leaders, mostly community nurses, who positively influenced professional development. Most nursing assistants felt that they needed to take initiative to learn new things. They generally felt supported and given sufficient space to do so:

> *“When you notice there is a need for it [advance care planning], I think that if I discuss it with the nurse, I can make an extra visit. (…) That way, you build a relationship of trust, and you can ask them: How are things going? What are you struggling with? What do you need? I feel that, within the team, we are given the space to do that*..*”* (P11)

Two participants mentioned hierarchy or dysfunctional team collaborations. They stopped their attempts to participate in additional tasks, also specifically in advance care planning:

> *“Things are not going very well in this team and there are a few colleagues who are in charge, and they just want to hold all the strings. [*…*] I do have something to say about this, but they pretty much have their own ideas about it* [advance care planning].*” (P09)*

These nursing assistants perceived their work environment and job satisfaction more negatively and also considered to leave their job.

#### Time pressure

Nursing assistants mentioned that a high workload and administrative tasks resulted in time pressure during advance care planning conversations. However, almost all nursing assistants mentioned the importance of making time for this:

> *“Sometimes there is a lack of time, but that is of course everywhere in healthcare. But you can also plan that, I think. There are always times during the week when it is less busy and you can schedule half an hour, so to speak, to have a quick chat or longer, where necessary. So I also think it is a mindset that you just have to deal with it differently*.*”* (P15)

In community care, nursing assistants mentioned that planning time for advance care planning could be challenging. Time for care moments was calculated beforehand, which often did not correspond to the clients’ care needs, especially when advance care planning was discussed. This required them to plan ahead or be creative in finding moments to initiate discussions about advance care planning:

> *“I also choose my moments when I stay for a cup of coffee with someone, and my breaks are always with the clients. (…) I choose a client where I notice that they need it at that moment*.*” (P11)*

### Motivation

#### Personal factors as (de)motivator

Nursing assistants’ main motivation to be involved in advance care planning was to improve the quality of life of clients in the last phase of life:

> *“Well, I think it is very important that in the last phase of life, things go the way someone wants. I think that is very important, that the person feels as if he is in control. (…) Yes, you know, that’s the only thing you can do for someone, right? To ensure that the last part goes well*.*”* (P13)

Some nursing assistants sought variety in their work by additional challenges and learning opportunities. These ambitions contributed to nursing assistants’ motivation to participate in advance care planning:

> *“I would like to make the care plan with the client, but then I would probably be one of a few nursing assistants. That’s just one thing that I really enjoy and maybe because I have this eagerness to continue learning [*…*] I just know that I think I can do more than what I’m showing now*.*” (P11)*

However, some nursing assistants preferred no work-related burdensome tasks due to personal circumstances, such as being an informal caregiver themselves. Furthermore, barriers in the performance of advance care planning, such as in interprofessional collaboration or in the communication with clients, were perceived as a demotivator:

> *“If a conversation* [about advance care planning] *is not progressing. That could really demotivate me from getting the right thing out. That you don’t get any further in a conversation and that you cannot get things clear. Then you stop the conversation, have to fill things in yourself or you end up with the family again*.*”* (P15)

#### The influence of team culture

The majority of participants reported that a positive team collaboration and support by colleagues were motivators to develop themselves and others in advance care planning:

> *“Some people need to be taken by the hand to watch, then they can become very good at it. (…). If you have motivated people, you also motivate others faster, so that is important*.*”* (P03)

In contrast, the two nursing assistants that experienced a negative team culture indicated that this influenced their opportunities to contribute to advance care planning, e.g., because they were not challenged or felt insufficiently included in the team:

> *“So I see myself play a larger role in it [advance care planning conversations], but that would not work in the place where I work now. I cannot do my job properly, and I cannot have these conversations. So what is important is a safe work environment so that you, as a healthcare professional, feel safe in the team. To have this kind of conversations, that you are taken seriously*.*”* (P05)

### Discussion

#### Main findings

This study showed that nursing assistants had varying roles in advance care planning. Their capability was influenced by their current role in advance care planning and their perceived knowledge and skills. Nursing assistants’ opportunities were shaped by their trust-based relationship with clients and relatives, possibilities within the care team and time pressure. Furthermore, personal factors and the team culture influenced nursing assistants’ motivation to be involved and professionally develop themselves in advance care planning. Several points of attention were identified that need to be considered regarding their role in advance care planning: 1) structural involvement in advance care planning, 2) fostering a supportive work environment and 3) empowerment.

#### What this study adds

##### Structural involvement in advance care planning

Our results demonstrated that large practice variation exists regarding the role of nursing assistants in advance care planning. This is in line with previous research describing the lack of clarity regarding the roles and responsibilities of all involved professionals in advance care planning, also including nursing assistants.^12, 24, 25^ Nursing assistants emphasized their added value in care for older persons in the last phase of life. Their regular involvement and trust-based relationship can contribute to important insights regarding palliative care needs and preferences beyond medical treatment preferences, such as (more psychosocial) life goals and wishes that are important for older persons’ quality of life. This is in line with previous research that described the added value of nursing professionals to look holistically at clients’ needs.^17, 25^

Previous research largely elaborated on the lack of knowledge and skills of healthcare professionals regarding advance care planning.^12, 15, 16, 26^ Nursing assistants in our study also indicated the need for additional education to support their structural involvement. For future nursing assistants, this topic should be an explicit part of their regular training. However, the educational preparation of nursing assistants differs internationally.^10, 27^ While in most European countries, programs last between one and three years and combine theory with practice, in some countries training is primarily workplace-based, with few or no formal educational requirements. These differences influence the extent to which nursing assistants can be optimally involved in advance planning, even though they all have an important role in signaling care needs, including palliative care needs.

##### Fostering a supportive work environment

Our results demonstrated that the team culture and the presence of a team leader, such as a registered nurse, can either facilitate or hinder nursing assistants’ opportunities and motivation to engage in advance care planning. Several nursing assistants in our study reported a negative work environment. These participants reported that they did not feel recognized or sufficiently challenged, which resulted in some nursing assistants intending to leave their jobs. Previous research emphasized that the work environment influences the quality of care, work engagement, job satisfaction and retention of nursing professionals.^28-32^ The retention of healthcare professionals is essential to meet the increasing and changing care demands of older persons. Healthcare organisations therefore need to critically look at the workplace support and division of tasks of healthcare professionals with different qualifications and expertise’s. Because palliative care, including advance care planning, relies on interprofessional collaboration, the role of nursing assistants also deserves explicit attention.

In the Netherlands, a professional profile describing the tasks and responsibilities of nursing assistants specialized in palliative care (providing palliative care for ≥50% of their working time) has been published recently by The Dutch Association for Nurses and Nursing Assistants (V&VN). ^33^ This profile aims to clarify their role for professionals, employers, and educators. Although not all nursing assistants need to be specialized in palliative care, such a profile may serve as a valuable starting point for discussing and defining their role and responsibility in practice. More broadly, healthcare organisations could benefit from reflecting on how to make optimal use of all professionals’ expertise. This may contribute to a more efficient and meaningful work process for advance care planning, while also enhancing job satisfaction and professional development among nursing assistants.

##### Empowerment

This study observed that nursing assistants sometimes spoke negatively or expressed doubt about their own capabilities (*“I am just a nursing assistant”*) which inhibit them from collaborating effectively with other disciplines in advance care planning, such as the responsible physician. Several studies showed that this is among others caused by other healthcare professionals who do not always know nursing assistants’ expertise and treat them as inferior.^8, 34^ However, nursing assistants also unconsciously position themselves as inferior when they collaborate with other professionals (such as managers or physicians), among others because of previous negative experiences or a lack of skills to share their perspectives.^35^ Previous research showed that self-efficacy was a more important predictor for nurses to engage in advance care planning as compared to knowledge.^14^ The empowerment of nursing assistants in advance care planning therefore seems to be crucial to become a more equal partner in collaboration with other involved professionals.^12^ This might contribute to the timely initiation of advance care planning and to appropriate palliative care for older persons, but also to nursing assistants’ job satisfaction and their sense of feeling valued in their work.^36, 37^

#### Strengths and limitations

This study is one of the first qualitative studies examining the perspectives of nursing assistants in the Netherlands regarding their role in advance care planning. The Capability, Opportunity, Motivation Behavioral model offered important insights regarding nursing into these perceptions, and its well-validated constructs support the study’s overall validity.^38, 39^

Some limitations need to be considered. All included participants were female, and the mean age was 52 years. However, this is largely representative of the current population of Dutch healthcare providers (mostly nursing professionals) of which 91% is female and nearly 31% ≥ 55 years.^40^ We did not purposively select nursing assistants from different ethnic backgrounds. Most of the respondents were of Dutch origin, while nursing assistants in the Netherlands are from many different origins. This study provided important first insights on their perspectives on advance care planning as their role is currently often overlooked, also in research. As culture and religion influence the perspectives on advance care planning, further research will specifically examine the perspectives of nursing assistants from different origins.

## Conclusion

This study found several barriers and facilitators affecting nursing assistants’ role in advance care planning for older persons. Structurally involving nursing assistants in advance care planning, supporting their work environment, and empowering them can foster equal collaboration with other healthcare professionals. This might contribute to the timely initiation of advance care planning and palliative care for older persons.

## Supporting information

Supplementary file 1. Description of advance care planning in the Netherlands

## Data Availability

Data relating to this research project is available upon request by contacting the corresponding author

## Declarations

### Authorship

PJ, OB and MvR contributed to the concept and design of the study. PJ, AB and OB conducted the qualitative interviews, analyzed the study data and drafted the original manuscript, under supervision of MvR. CL, BB and MvR made substantial contributions to the manuscript and revised it for important intellectual content. All authors provided critical feedback to several drafts of the manuscript.

### Funding

This work was supported by the Netherlands Organisation for Health Research and Development (ZonMw) grant number 80-86300-98-118 to MvR. The sponsors had no role in study design, data collection and analysis, and neither in the preparation or publication of the manuscript.

### Conflict of interest

The authors declare that there is no conflict of interest.

### Ethics and consent

The Medical Ethics Review Committee at Amsterdam University Medical Center approved our interviews (approval: 2022.0868) on January 25, 2023. Respondents gave written consent for review and signature before starting interviews.

## Acknowledgements

We are grateful for all participating certified nursing assistants in this study.

## Notes

### Competing Interest Statement

The authors have declared no competing interest.

## References

1. Russell AE, Denny R, Lee PG, et al. Palliative care considerations in frail older adults. Ann Palliat Med 2024; 13: 976–990. 20240606. DOI: 10.21037/apm-23-559.

2. Ganz FD, Roeh K, Eid M, et al. The need for palliative and support care services for heart failure patients in the community. Eur J Cardiovasc Nurs 2021; 20: 138–146. DOI: 10.1177/1474515120951970.

3. Jabbarian LJ, Zwakman M, van der Heide A, et al. Advance care planning for patients with chronic respiratory diseases: a systematic review of preferences and practices. Thorax 2018; 73: 222–230. 20171106. DOI: 10.1136/thoraxjnl-2016-209806.

4. Van den Block L, de Nooijer K, Pautex S, et al. A European Association for Palliative Care White Paper defining an integrative palliative, geriatric, and rehabilitative approach to care and support for older people living with frailty and their family carers: a 28-country Delphi study and recommendations. EClinicalMedicine 2025; 87: 103403. 20250812. DOI: 10.1016/j.eclinm.2025.103403.

5. Rietjens JAC, Sudore RL, Connolly M, et al. Definition and recommendations for advance care planning: an international consensus supported by the European Association for Palliative Care. The Lancet Oncology 2017; 18: e543–e551. DOI: 10.1016/S1470-2045(17)30582-X.

6. Bekker YAC, Suntjens AF, Engels Y, et al. Advance care planning in primary care: a retrospective medical record study among patients with different illness trajectories. BMC Palliat Care 2022; 21: 21. 20220214. DOI: 10.1186/s12904-022-00907-6.

7. Glaudemans JJ, Van Charante EPM and Willems DL. Advance care planning in primary care, only for severely ill patients? A structured review. Family Practice 2015; 32: 16–26. DOI: 10.1093/FAMPRA/CMU074.

8. van Wieringen M, Wendelgelst R and Gobbens RJJ. ‘They’re not doing too much are they?’ How the socialization of registered nurses perpetuates status differences with certified nursing assistants: A qualitative study. Nurse Educ Today 2023; 131: 105984. 20231011. DOI: 10.1016/j.nedt.2023.105984.

9. van Kraaij J, Veenstra M, Stalpers D, et al. Uniformity along the way: A scoping review on characteristics of nurse education programs worldwide. Nurse Educ Today 2023; 120: 105646. 20221119. DOI: 10.1016/j.nedt.2022.105646.

10. Jackson J, Gadimova F and Epko S. A nurse by any other name? An international comparison of nomenclature and regulation of healthcare assistants. Int J Nurs Stud Adv 2024; 6: 100200. 20240416. DOI: 10.1016/j.ijnsa.2024.100200.

11. Van Wieringen M, Kee K, Nies HLG, et al. Verzorgenden IG in beeld: Samen werken aan een duidelijke stem en betere positie voor de beroepsgroep. [Certified nursing assistants in the picture: Working together towards a clear voice and better position for the professional group]. Research Report. 2021.

12. Punia H, Kaasalainen S, Ploeg J, et al. Exploring the Role of Nurses in Advance Care Planning Within Long-Term Care Homes: A Qualitative Study. SAGE Open Nurs 2024; 10: 23779608241249335. 20240429. DOI: 10.1177/23779608241249335.

13. van Doorne I, Mokkenstorm K, Willems DL, et al. The perspectives of in-hospital healthcare professionals on the timing and collaboration in advance care planning: A survey study. Heliyon 2023; 9: e14772. 20230322. DOI: 10.1016/j.heliyon.2023.e14772.

14. Gilissen J, Pivodic L, Wendrich-van Dael A, et al. Nurses’ self-efficacy, rather than their knowledge, is associated with their engagement in advance care planning in nursing homes: A survey study. Palliat Med 2020; 34: 917–924. 20200508. DOI: 10.1177/0269216320916158.

15. Gilissen J, Pivodic L, Smets T, et al. Preconditions for successful advance care planning in nursing homes: A systematic review. Int J Nurs Stud 2017; 66: 47–59. 20161208. DOI: 10.1016/j.ijnurstu.2016.12.003.

16. Ke LS, Huang X, O’Connor M, et al. Nurses’ views regarding implementing advance care planning for older people: a systematic review and synthesis of qualitative studies. J Clin Nurs 2015; 24: 2057– 2073. 20150504. DOI: 10.1111/jocn.12853.

17. Jepma P, Eijk R, Bos AAE, et al. Feasibility of a new transmural care pathway for advance care planning for older persons: A qualitative study into community care registered nurses’ perspectives. Int J Nurs Stud Adv 2024; 7: 100264. 20241108. DOI: 10.1016/j.ijnsa.2024.100264.

18. Kwak J, Jamal A, Jones B, et al. An Interprofessional Approach to Advance Care Planning. Am J Hosp Palliat Care 2022; 39: 321–331. 20210607. DOI: 10.1177/10499091211019316.

19. Sandelowski M. Whatever happened to qualitative description? Res Nurs Health 2000; 23: 334– 340. DOI: 10.1002/1098-240x(200008)23:4<334::aid-nur9>3.0.co;2-g.

20. Tong A, Sainsbury P and Craig J. Consolidated criteria for reporting qualitative research (COREQ): a 32-item checklist for interviews and focus groups. International Journal for Quality in Health Care 2007; 19: 349–357. DOI: 10.1093/INTQHC/MZM042.

21. Malterud K, Siersma VD and Guassora AD. Sample Size in Qualitative Interview Studies: Guided by Information Power. Qual Health Res 2016; 26: 1753–1760. 20160710. DOI: 10.1177/1049732315617444.

22. Michie S, van Stralen MM and West R. The behaviour change wheel: a new method for characterising and designing behaviour change interventions. Implement Sci 2011; 6: 42. 20110423. DOI: 10.1186/1748-5908-6-42.

23. Elo S and Kyngäs H. The qualitative content analysis process. Journal of Advanced Nursing 2008; 62: 107–115. DOI: 10.1111/j.1365-2648.2007.04569.x.

24. Poveda-Moral SA-O, Falcó-Pegueroles AA-O, Ballesteros-Silva MA-O, et al. Barriers to Advance Care Planning Implementation in Health care: An Umbrella Review with Implications for Evidence-Based Practice. Worldviews Evid Based Nurs 2021; 18: 254–263.

25. Bolt SR, van der Steen JT, Schols JMGA, et al. [The nurse’s role in the process of advance care planning]. Tijdschr Gerontol Geriatr 2021; 52. DOI: doi: 10.36613/tgg.1875-6832/2021.01.0.

26. Blackwood DH, Walker D, Mythen MG, et al. Barriers to advance care planning with patients as perceived by nurses and other healthcare professionals: A systematic review. J Clin Nurs 2019; 28: 4276– 4297.

27. Kroezen M, Schäfer W, Sermeus W, et al. Healthcare assistants in EU Member States: An overview. Health Policy 2018; 122: 1109–1117. 20180720. DOI: 10.1016/j.healthpol.2018.07.004.

28. Lamiani G, Borghi L and Argentero P. When healthcare professionals cannot do the right thing: A systematic review of moral distress and its correlates. J Health Psychol 2017; 22: 51–67. 20160710. DOI: 10.1177/1359105315595120.

29. Keyko K, Cummings GG, Yonge O, et al. Work engagement in professional nursing practice: A systematic review. Int J Nurs Stud 2016; 61: 142–164. 20160608. DOI: 10.1016/j.ijnurstu.2016.06.003.

30. Aloisio LD, Gifford WA, McGilton KS, et al. Factors Associated With Nurses’ Job Satisfaction In Residential Long-term Care: The Importance of Organizational Context. J Am Med Dir Assoc 2019; 20: 1611–1616.e1614. 20190806. DOI: 10.1016/j.jamda.2019.06.020.

31. Bloemhof J, Knol J, Van Rijn M, et al. Organising Nurse Work Environments: (Reshaped) Roles of Nursing Teams-A Qualitative Descriptive Study. J Adv Nurs 2025 20250728. DOI: 10.1111/jan.70093.

32. Patynowska KA, Maun E, Fantoni ER, et al. Workplace support, wellbeing and intention to leave among lone working healthcare assistants providing palliative and end-of-life care in the community: A mixed methods study. Palliat Med 2025: 2692163251395576. 20251206. DOI: 10.1177/02692163251395576.

33. The Dutch Association for Nurses and Nursing Assistants VV. Profiel gespecialiseerd verzorgenden IG palliative zorg. [Profile specialized nursing assistants palliative care]. 2024.

34. Zysberg L, Band-Winterstein T, Doron I, et al. The health care aide position in nursing homes: A comparative survey of nurses’ and aides’ perceptions. Int J Nurs Stud 2019; 94: 98–106. 20190318. DOI: 10.1016/j.ijnurstu.2019.03.007.

35. Kee K, van Wieringen M and Beersma B. The relational road to voice: how members of a low-status occupational group can develop voice behavior that transcends hierarchical levels. Journal of Professions and Organization 2021; 8: 253–272. DOI: 10.1093/jpo/joab011.

36. Travers JL, Schroeder K, Norful AA, et al. The influence of empowered work environments on the psychological experiences of nursing assistants during COVID-19: a qualitative study. BMC Nurs 2020; 19: 98. 20201016. DOI: 10.1186/s12912-020-00489-9.

37. Wonder AH, Martin EK and Jackson K. Supporting and Empowering Direct-Care Nurses to Promote EBP: An Example of Evidence-Based Policy Development, Education, and Practice Change. Worldviews Evid Based Nurs 2017; 14: 336–338. DOI: 10.1111/wvn.12239.

38. Luo C, Yang C, Yuan R, et al. Barriers and facilitators to technology acceptance of socially assistive robots in older adults - A qualitative study based on the capability, opportunity, and motivation behavior model (COM-B) and stakeholder perspectives. Geriatr Nurs 2024; 58: 162–170. 20240529. DOI: 10.1016/j.gerinurse.2024.05.025.

39. Den Hamer-Jordaan G, Groenendijk-Van Woudenbergh GJ, Haveman-Nies A, et al. Factors associated with dietary behaviour change support in patients: A qualitative study among community nurses. J Adv Nurs 2024; 80: 500–509. 20230730. DOI: 10.1111/jan.15808.

40. Statistics Netherlands. Labour market profile of care and welfare in 2022: Healthcare workers. 2023., https://www.cbs.nl/nl-nl/longread/statistische-trends/2023/arbeidsmarktprofiel-van-zorg-en-welzijn-in-2022/3-zorgmedewerkers (accessed 09-29 2024).

