## Supplementary file 1. Description of advance care planning in the Netherlands for "Certified nursing assistants’ perspectives on their role in advance care planning for older persons: a qualitative study"

In the Netherlands, palliative care is provided according to the generalist plus specialist model.[1-3] In this model, core elements of palliative care, such as aligning care and treatment to patients' preferences', can be provided by any healthcare professional involved including physicians, nurses and paramedics. In primary care, the general practitioner is often the primary responsible physician as they are often closely involved in the care and treatment throughout the life span of their patients and could therefore have an important role in continuity of care, including care at in the last phase of life.[3] Also medical specialists from other settings could be involved or could be the responsible physician, such as the medical specialist in the hospital setting or the elderly care physician in a nursing home. The primary responsible physician is responsible for initiating advance care planning conversations, the registration of advance directives in the medical file (e.g. regarding resuscitation), and the follow-up and evaluations of agreements over time. In line with the generalist plus specialist model, palliative care specialists could be consulted in complex situations to work in collaboration with the responsible physician.[2]

Advance care planning, in terms of medical preferences is thus primarily the responsibility of the primary physician.[3] However, according to the definition of advance care planning, it addresses individuals' concerns across the physical, psychological, social, and spiritual domains.[4] This makes advance care planning an interprofessional task with a need for collaboration with other involved healthcare professionals involved in palliative care (such as nursing professionals). In 2020, a national educational programme in the Netherlands created the Palliative Care Competence Framework for healthcare professionals in different healthcare settings, describing roles and responsibilities for certified nursing assistants, registered nurses and physicians up to level 8 of the Dutch National Qualifications Framework (NLQF)[5], referring to the level of the medical specialist (for example, in internal medicine, oncology, cardiology). For each NLQF level, the necessary individual palliative care competences are being described in detail. Furthermore, the educational programme also contributes to more awareness for interprofessional collaboration in palliative care in different care settings (e.g. hospital, nursing home and community care), including the role of nursing professionals in advance care planning conversations. For example, they can have preparatory conversations with older persons and their relatives to help address (non-medical) life goals and wishes that are important for older persons' quality of life.[6] Subsequently, older persons can formally discuss their treatment preferences and limitations with the responsible physician, who is also responsible for the documentation of agreements in the medical record and for the regular evaluation.[1]

Certified nursing assistants are closely involved in the daily care of older persons, and therefore have an important role in the observation of care preferences and key aspects regarding their clients' quality of life.[7] his can provide important information for involved registered nurses and the responsible physician regarding potential palliative care needs. Recently, The Dutch association for nurses and nursing assistants (V&VN), department of palliative care has published a national professional profile specifically for certified nursing assistants specialized in palliative care (providing palliative care ≥ 50% of work time).[8] Competences include among others the identification of the palliative phase, the observation of palliative care needs and communication skills, all of which are also of great importance for advance care planning. This might contribute to increasing awareness of the potential role of Dutch certified nursing assistants in advance care planning and can contribute to clarity on their added value in interprofessional palliative care.

**References**


1. Boddaert MDJ DJ, Dijxhoorn F, Bijkerk M. *Quality Framework for Palliative Care in the Netherlands [Kwaliteitskader palliatieve zorg Nederland]*. IKNL/Palliactief, 2017. https://palliaweb.nl/richtlijnen-palliatieve-zorg/richtlijn/kwaliteitskader-palliatieve-zorg-nederland.
2. Quill TE and Abernethy AP. Generalist plus Specialist Palliative Care — Creating a More Sustainable Model. *New England Journal of Medicine* 2013; 368: 1173–1175. DOI: 10.1056/NEJMp1215620.
3. van der Steen JT, Engels Y, Touwen DP, et al. Advance Care Planning in the Netherlands. *Z Evid Fortbild Qual Gesundhwes* 2023; 180: 133–138. 20230721. DOI: 10.1016/j.zefq.2023.06.003.
4. Rietjens JAC, Sudore RL, Connolly M, et al. Definition and recommendations for advance care planning: an international consensus supported by the European Association for Palliative Care. *Lancet Oncol* 2017; 18: e543–e551. DOI: 10.1016/s1470-2045(17)30582-x.
5. The Dutch Qualifications Framework (NLQF), https://www.nlqf.nl/english (accessed October 14 2025).
6. Jepma P, Eijk R, Bos AAE, et al. Feasibility of a new transmural care pathway for advance care planning for older persons: A qualitative study into community care registered nurses' perspectives. *Int J Nurs Stud Adv* 2024; 7: 100264. 20241108. DOI: 10.1016/j.ijnsa.2024.100264.
7. Van Wieringen M, Kee K, Nies HLG, et al. Verzorgenden IG in beeld: Samen werken aan een duidelijke stem en betere positie voor de beroepsgroep. [Certified nursing assistants in the picture: Working together towards a clear voice and better position for the professional group]. Research Report. 2021.
8. The Dutch Association for Nurses and Nursing Assistants VV. Profiel gespecialiseerd verzorgenden IG palliative zorg. [Profile specialized nursing assistants palliative care]. 2024.
